# Toward Understanding Genetic Risk of Burnout through Genome-wide Associations of Cynicism and Cynical Distrust

**DOI:** 10.1101/2023.05.16.23289156

**Authors:** Richard R Chapleau

## Abstract

Cynicism is an attitude that people are motivated primarily by self-interest, and often manifests alongside emotions like contempt, anger, and hostility. Cynicism can impact all aspects of an individual’s life: in the workplace it is a critical component of burnout, while at home it often leads to marital dissatisfaction and increased family conflicts. Using data from 2,317 participants in the CARDIA Cohort study, we performed a genome-wide association study (GWAS) to identify genetic variants related to cynicism and the cynical distrust sub-trait. We performed two GWAS tests for each trait, one without controlling for population stratification and a second where we considered possible ancestry-related stratification using principal component analysis within plink 1.9. We also tested multiple genetic inheritance models. Finally, we performed pathway analysis using the STRINGdb web resource to identify meaningful relationships. From our various approaches, we identified 12 variants associated with high cynicism in an additive model and 258 in a genotypic model. We also identified 24 and 275 variants associated with cynical distrust using the respective models. Our network analysis revealed clear pathways for how cynicism can lead to health outcomes like cardiovascular disease, adrenal disorders, neurological disorders, and even addiction. Specifically, genes involved in 30 distinct KEGG pathways were related to cynicism and clear connections can be made between cynicism and cardiomyopathies, Cushing syndrome, Alzheimer’s disease, and amphetamine addiction. Each of the outcomes are supported by gene association with at least 3 intermediate pathways. Our results provide the first objective evidence supporting the observational literature connections to cardiovascular disease, Alzheimer’s disease, and addiction. To the best of our knowledge, this is the first evidence presenting a relationship between cynicism and Cushing’s syndrome, demonstrating the power of population and statistical genetics.

## INTRODUCTION

Cynicism is an attitude of distrusting the motives or sincerity of people and their actions, and a belief that people are motivated purely by self-interest [1]. Cynicism can also imply a pessimistic outlook on the future or a sarcastic tone. It can also mean an inclination to question whether something will happen or whether it is worthwhile.

Cynicism can have negative effects on health and mortality [2]. For example, middle-age and older people who are highly stressed, have depression or who are perhaps even just cynical may be at increased risk of stroke. In a study of more than 6,700 healthy adults ages 45 to 84 [3], researchers found that people with high levels of cynicism were more than twice as likely to have a stroke compared with their less cynical counterparts. The results of the study held even when researchers accounted for known risk factors of stroke, including age, race, sex, and health behaviors. The findings suggest that psychological well-being, which has already been linked to heart health, also plays a role in stroke risk.

Cynical hostility may also cause an increased risk of developing cardiovascular disease (CVD) [4]. Results from 196 participants in a stress test alongside a modified version of the Cook-Medley Hostility Scale (CMHS) suggest that stress responses had no relationship to emotional or behavioral hostility, however cynicism was associated with decreased cardiovascular response and reactivity. These results imply that the increased risk of hostility is likely due to heightened physiological arousal to psychological stress, which can strain the cardiovascular system over time. With cynical hostility, a person continues to react to stressful circumstances with a similar intensity level, regardless of the amount and frequency of similar stress exposures. Consistent arousal of this nature causes a strain on the cardiovascular system over time.

Another study found that people with high levels of cynical distrust (CD) may be more likely to develop dementia [5]. After adjusting for confounders, higher CD was associated with increased risk of dementia. In a longitudinal study following 622 people aged 65 to 79 over 10 years, the likelihood of individuals with high levels of CD developing dementia was more than triple the likelihood of those individuals with low CD levels (relative risk = 3.13; 95% confidence interval 1.15-8.55). The mechanisms by which CD and cynicism lead to increased neurological decline are unclear, but the researchers hypothesize that cynical people may have higher levels of inflammation or different levels of stress-related hormones.

To date, genetic risk markers for cynicism have only recently been reported [6], despite the inclusion of the CMHS in large scale genomics studies [7,8]. Genetic risk markers for cynicism may be able to serve as additional pieces of information regarding life-long physical and mental health outcomes. Here we report the results of a genome-wide association study (GWAS) concerning cynicism and CD. In addition to genetic associations, we also report the results of gene network analyses and interpret pathways through which cynicism may exert its impact on CVD, neurological disorders, and other health outcomes.

## MATERIALS AND METHODS

As with the prior study [6], this study was reviewed and approved by WCG IRB (Study number 1332892) and was performed in accordance with the Helsinki declaration. Data were obtained from the National Institutes of Health’s Database of Genotypes and Phenotypes (dbGAP) using the Coronary Artery Risk Development in Young Adults (CARDIA) Study (dbGAP study accession phs000285) [8]. As this is a genetic association study, we followed the STREGA guidelines available from the EQUATOR network [9].

### Phenotype and Genotype Data pre-processing

Cynicism and CD were measured as 12- and 8-item subscales of the CMHS, respectively [10-12]. Scores for each trait were calculated as a mean of the responses. We also defined binary variables for high cynicism and high CD as individuals with scores above the medians [13,14]. Missing data were replaced with the mean. Affymetrix Human SNP-6 microarray genotype data were obtained from two CARDIA sub-studies (dbGAP accession numbers phg000092.v2 and phg000098.v2). Binary plink file sets were merged and filtered for autosomal genotypes with less than 10% missing genotype calls (sample or locus), a minor allele frequencies threshold was set at 1%, and a Hardy-Weinberg equilibrium threshold of .0001. The total sample size was 2317. Genetic pre-processing was performed using plink1.9 and phenotype processing was performed in R version 4.2.0.

### Unadjusted GWAS

We conducted GWAS using plink 1.9 evaluating the total scores (not scores for the questionnaire responses) and high/low status. We performed four association analyses (using the --linear recessive/genotypic/hethom/dominant flags for plink, as appropriate) for each outcome testing a total of six inheritance models (additive, dominant, recessive, genotypic, dominance deviation, heterozygotic, and homozygotic) using linear or logistic regressions, as appropriate and determined autonomously by plink during execution.

### Population-stratification adjusted GWAS

To account for the possibility of population stratification, we performed principal component analysis (PCA) in plink. We used the resulting eigenvector file as the covariate input file and included the first four principal components (PC) as covariates in subsequent association tests. We performed the same four association analyses as for the unadjusted GWAS testing the same six inheritance models.

### Gene annotations and network analysis

The resulting significant associations were annotated with gene and variant identifiers using either SNPtracker [15] or FAVOR v2.0 [16]. The dbSNP Batch Query provided annotations based upon the GRCh38, dbSNP build 155 and the FAVOR annotations are built on GRCh38 including the Trans-Omics for Precision Medicine (TOPMed) Freeze 8 BRAVO variant set [17]. Interaction networks were predicted using web-interface of STRING (www.string-db.org) [18] with a minimum association score of 0.150, no more than 5 first shell interactions added when the query protein list had fewer than 20 proteins, and no second shell interaction additions permitted regardless of number of query proteins.

## RESULTS

The characteristics of the dataset have been described previously. Briefly, the median cynicism score was 6 and there were 1,228 high cynicism samples. For CD, the median was 3 and there were 1,163 high CD samples. Gender distribution favored females (58%). Variables were non-normally distributed (Shapiro-Wilk’s *W* = 0.935 and 0.970 for cynicism and CD scores, respectively; and 0.615 and 0.622 for high cynicism and high CD, respectively), skewed (0.118 and 0.373 for cynicism and CD, respectively), and platykurtic (1.01 and 2.18 for cynicism and CD, respectively).

### Unadjusted GWAS Results

Consistent with our prior results not accounting for population stratification, we observed a very large number of nominally significant associations (P < 5×10^−8^) to all four traits (Figure 1, white columns). Using the additive model of inheritance, we identified between 4,800 and 13,800 significantly associated variants depending upon the trait. Five of the other six inheritance models tested (dominant, recessive, genotypic, homozygous, heterozygous, and dominance deviation) yielded thousands of associated variants in each trait. Interestingly, the dominance deviation model did not result in many significant associations. The large number of significant associations may suggest an underlying population stratification artifact.

**Figure 1:**
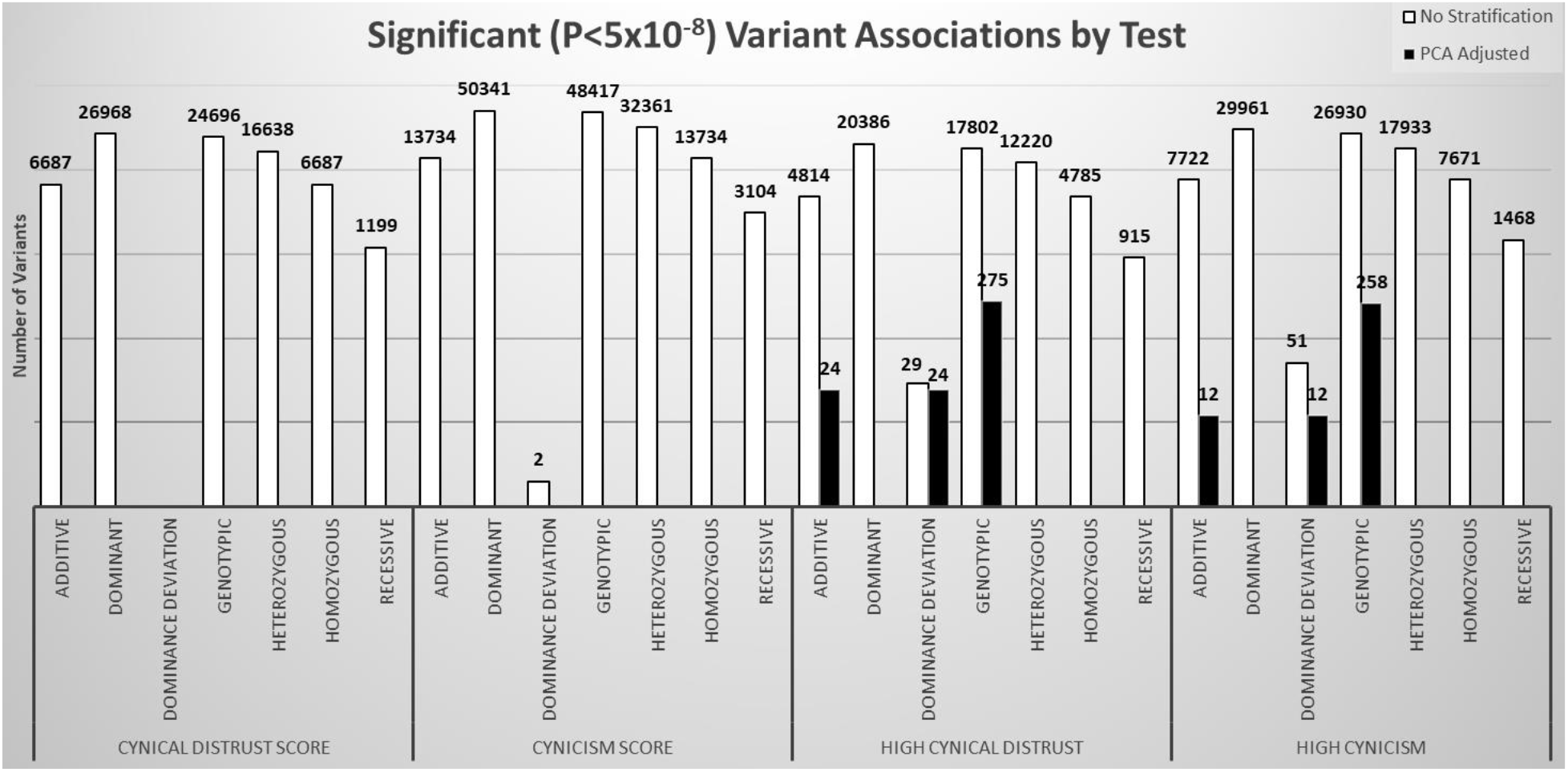
Count of GWAS significant (P<5×10^−8^) variants associated with cynical distrust sore, cynicism score, high cynical distrust, or high cynicism. There are many thousands of significantly associated variants for each trait in all models except dominance deviation when not accounting for population stratification (white columns). When stratification is accounted for by including principal components as covariates (black columns), all associations disappear for trait scores and the number of associations for the binary traits is much lower.

### PCA-Adjusted GWAS Results

Our PCA analysis found that the first PC accounted for 74.9% of the variance in the dataset. Additionally, the first four PCs explained 81.3% of the variance and fitting a linear trend to the full dataset in a Scree plot (Supplementary Figure S1) has a lower goodness-of-fit value (R^2^ = 0.989) compared to fitting the first four PCs and the remaining 16 PCs (0.999 and 0.999, respectively). Therefore, we used the first four PCs to account for population stratification (Supplementary Figure S2 shows population dispersion by PC).

As with the unadjusted GWAS, we tested six inheritance models (Figure 1, black columns), but only observed significant associations with the traits when using additive, dominance deviation, and the genotypic inheritance models. We found 12 variants to be significantly associated (P < 5×10^−8^) with cynicism in the additive and dominance deviation models and 258 variants associated via the genotypic model (Figure 2). The complete list of variants is provided as supplementary material. In accordance with the CARDIA data use agreement, public posting of the complete summary statistics is not allowed and therefore we are only able to provide the effect sizes (odds ratios) and P-values of association for the significantly associated variants. Similarly, for CD we identified 24 significant variants in the additive and dominance deviation models and 275 variants in the genotypic model (Figure 3). All variants in the additive and dominance deviation models were the same. All 12 variants associated with cynicism and 24 variants associated with CD in the additive inheritance models were also identified in the genotypic models. None of the variants in the additive models were associated with both traits. However, 5 of the variants identified in the genotypic model were associated with both traits (chr3:72620060, chr4:166515714, chr11:109909301, chr16:27696779, and chr18:19171829).

**Figure 2:**
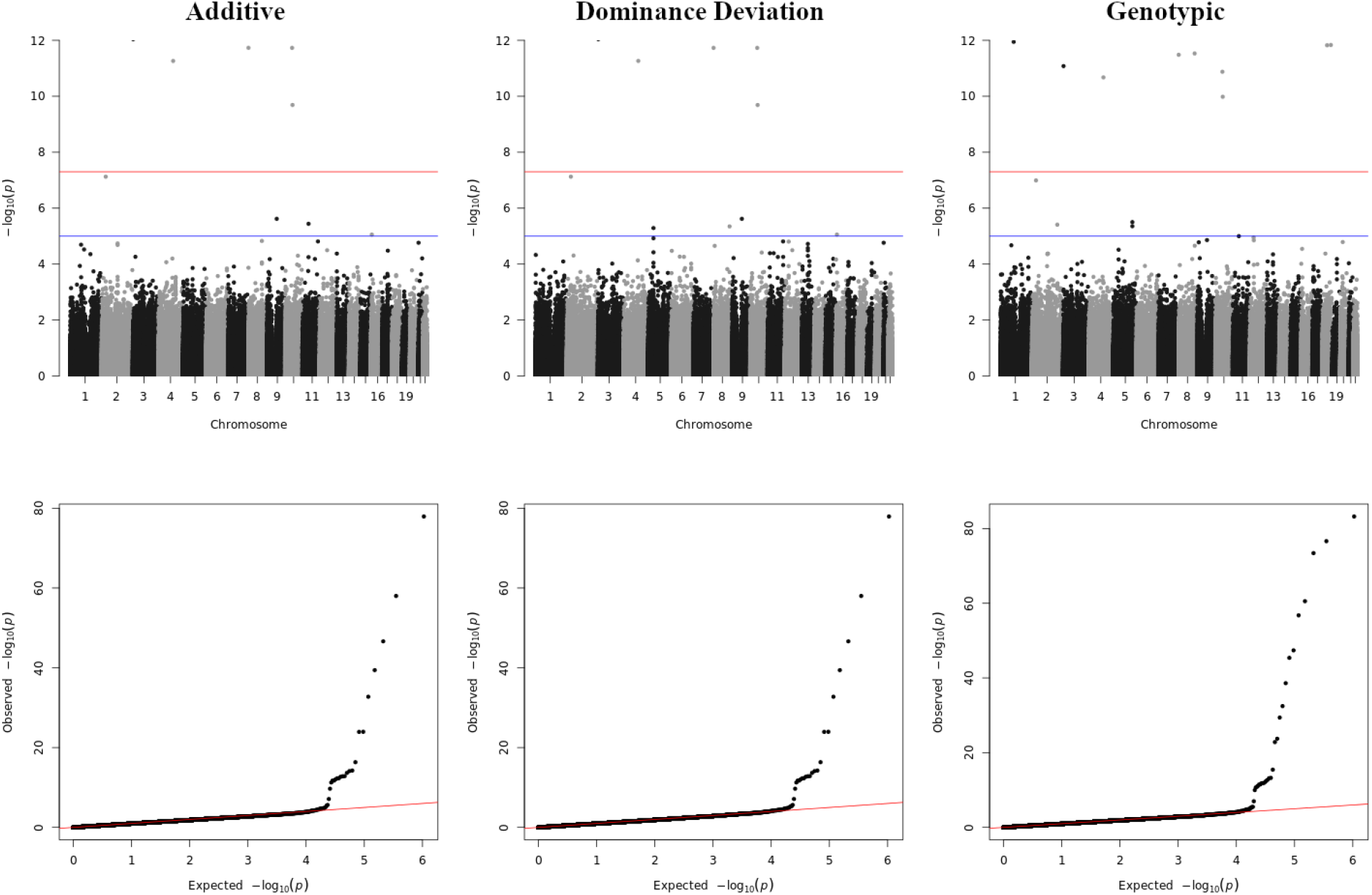
Manhattan and quartile-quartile plots for genetic associations with high cynicism.

**Figure 3:**
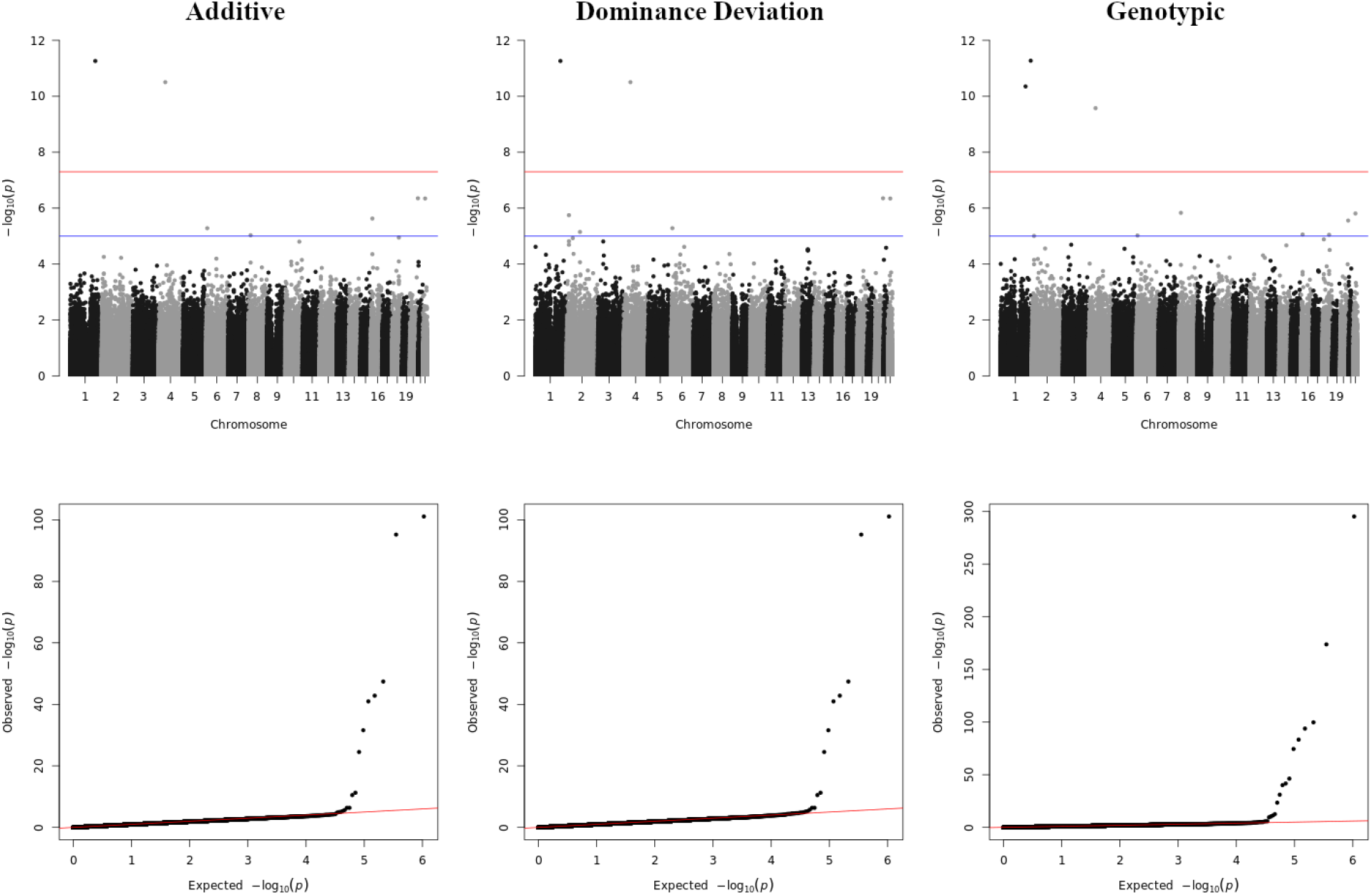
Manhattan and quartile-quartile plots for genetic associations with high cynical distrust.

### Network Analysis on PCA-Adjusted Significant SNPs

After annotating our significant variants for associated genes, we performed a network analysis using the STRINGdb web resource. The results from our additive models had fewer than 20 proteins to query so we included up to 5 first-order interacting proteins in the search criteria (Figure 4A and C for cynicism and CD, respectively). We did not include interacting proteins for the genes identified using the genotypic model (Figure 4B and D). The protein-protein interaction P-values reported by STRINGdb were significant for CD in the additive and genotypic models and for the genotypic model of cynicism (P = 0.013, 1.12×10^−10^, and <1.0×10^−16^, respectively). The additive model of cynicism, however, did not reach significance (P= 0.11) suggesting that the number of interactions observed did not differ from those expected of a random collection of genes. However, when we increased the confidence threshold of interactions from 0.15 to 0.40, all four trait-model pairs reached significance (P = 0.01, 0.002, 6.8×10^−8^, and 0.03, respectively). In addition to the network interaction diagrams, STRINGdb allows for searching multiple databases for pathways affected by genes in the query network. Our analysis revealed entries in 14 distinct databases, and collectively our networks accounted for 317 enriched terms across these databases (Table 1).

**Figure 4:**
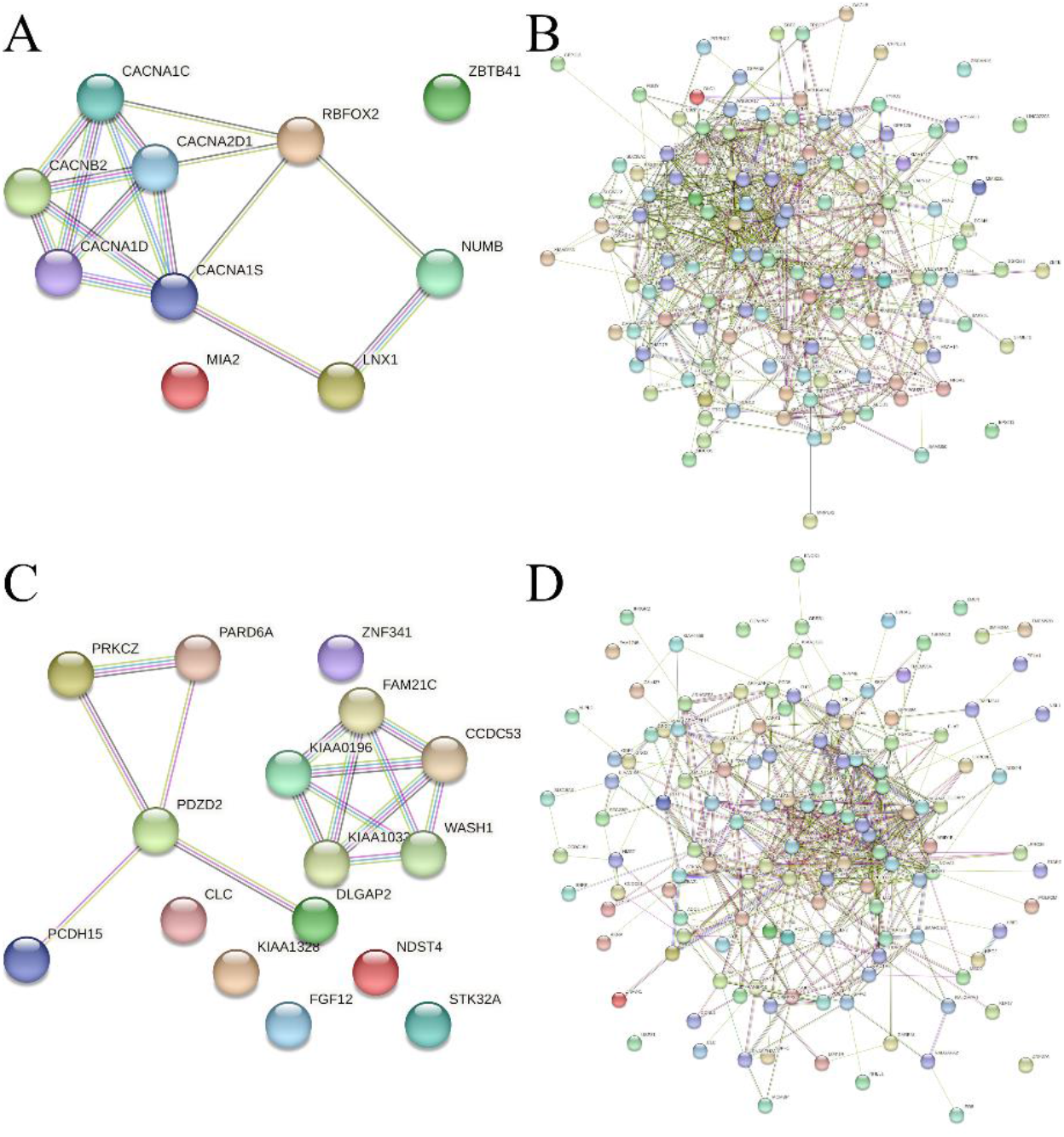
Network diagrams for genes significantly associated with cynicism (A and B) or cynical distrust (C and D). The additive models produced fewer nodes than 20 nodes (A and C) and therefore include 5 first-order network connection whereas the genotypic models (B and D) produced more than 20 nodes.

**Table 1:** Counts of enriched terms in STRINGdb pathway and network databases.

|  | CD | CD | Cynicism | Cynicism |
| --- | --- | --- | --- | --- |
|  | Additive | Genotypic | Additive | Genotypic |
| *Annotated Keywords (UniProt)* | 2 | 1 | 3 | 2 |
| *Biological Process (Gene Ontology)* | 0 | 0 | 26 | 0 |
| *Cellular Component (Gene Ontology)* | 3 | 0 | 3 | 0 |
| *Molecular Function (Gene Ontology)* | 0 | 0 | 5 | 0 |
| *Human Phenotype (Monarch)* | 0 | 72 | 13 | 119 |
| *KEGG Pathways* | 1 | 1 | 30 | 0 |
| *Protein Domains (Pfam)* | 0 | 2 | 5 | 0 |
| *Protein Domains & Features (InterPro)* | 0 | 0 | 7 | 0 |
| *Reactome Pathways* | 0 | 0 | 5 | 0 |
| *Subcellular localization (COMPARTMENTS)* | 1 | 0 | 1 | 0 |
| *Disease-gene associations (DISEASES)* | 0 | 0 | 1 | 0 |
| *Protein Domains (SMART)* | 1 | 1 | 1 | 0 |
| *Local network cluster (STRING)* | 2 | 0 | 4 | 0 |
| *WikiPathways* | 0 | 0 | 5 | 0 |
| *Total entries* | 10 | 77 | 109 | 121 |

## DISCUSSION

Our results from genetic analyses suggest that there was a high degree of population stratification in the CARDIA cohort. In our original two-stage approach, we had a discovery sample size of 1,852 participants after data cleaning. In the present study where we used all data collectively, we had 2,317 participants which is a net increase of more than 25%. In the prior study using an additive inheritance model, we identified nearly 29,000 candidate genetic variants associated with cynicism and more than 14,000 associated with CD. Without accounting for population stratification in our current study, we identified approximately half as many variants in the additive model (Figure 1). In total, 4,918 variants associated with cynicism in the original study were reproduced in the unadjusted association test here (17.0% reproduced associations) and 2,357 variants associated with CD were reproduced here (16.7%).

In contrast, when we accounted for population stratification by using the first four PCs, the number of associated variants dropped dramatically. In our PCA-adjusted analysis, we found only 12 significant associations with cynicism and 24 with CD, a meager 0.04% and 0.17% of the original study, respectively. None of the 12 cynicism-associated variants or the 24 CD-associated variants from the adjusted study were present in the original study list or the variant list from the current study’s unadjusted GWAS.

It is well known that misspecification of genetic models in GWAS is prone to increasing type II error rates, but also that multiple model testing can increase type I error [19-21]. As the effect sizes are expected to be small and the variants are expected to be common for these traits, we elected to test multiple inheritance models. All variants identified in the additive models were also identified in the dominance deviation models, indicating that there is no additional dominance of one allele over the other, so for clarity we only discuss the differences between the additive and genotypic models. The genotypic model makes no assumptions about the inheritance pattern, and therefore was expected to have a large number of variants, many of which are unlikely to replicate in more specific models or additional testing.

Our network analysis revealed a few interesting connections between cynicism and health outcomes. Cynicism has been previously shown to be a potential causal factor for cardiovascular diseases (CVD) [4, 22]. Our results here provide further support for these observations and provide a potential mechanism by which cynicism may lead to CVD. One of the networks identified from the pathway analysis was renin secretion (Kyoto Encyclopedia of Genes and Genomes, KEGG, pathway number: hsa04924). Renin secretion is involved in modulating three signaling pathways: calcium (hsa04020), cGMP-PKG (hsa04022) and cAMP (hsa04024). These three pathways then modulate the adrenergic signaling in cardiomyocytes (hsa04261) which controls cardiac and vascular smooth muscle contraction (hsa04260 and hsa04270, respectively). Changes in the contractility of cardiac and vascular smooth muscles can lead to hypertrophic (hsa05410), arrhythmogenic right ventricular (hsa05412) and dilated (hsa05414) cardiomyopathies. Importantly, these pathways are only named because we found a genetic variant involved in these pathways in our cynicism GWAS.

In addition to CVD, neurological disorders like Alzheimer’s disease (hsa05010) and dementia have been previously associated with cynicism [5, 23]. While our study did not find a significantly associated genetic variant with dementia, we did observe significant variants in genes associated with Alzheimer’s disease and a potential pathway to manifestation of the disease via insulin secretion (hsa04911) modifying calcium (hsa04020) and MAPK (hsa04010) signaling pathways eventually leading to Alzheimer’s disease [24].

Another disorder with observational evidence supporting a connection to cynicism is the development of amphetamine addiction (hsa05031) [25,26]. Our results suggest a shared pathway to chronic exposure of amphetamines through modified calcium (hsa04020) and cAMP (hsa04024) signaling at the glutamatergic (hsa04724) and dopaminergic (hsa04728) synapses.

Together with the prior evidence suggesting an increased risk of heroin addiction in highly cynical individuals [20], our results suggest that pain management alternatives to morphine would be better options for reducing addiction risk in populations with high occupational burnout such as medical care providers or military personnel.

Finally, we also identified one potentially rare disorder with no prior evidence supporting a connection. Cushing syndrome results from chronic exposure to excess glucocorticoids and typically occurs at a prevalence of approximately 40 cases per million person-years [27]. The usual lab tests for this disorder involve directly measuring salivary cortisol levels or response to corticosteroids. Here, we found variants associated with genes involved in cortisol and aldosterone synthesis and secretion (hsa04927 and hsa04925, respectively). One possible pathway to Cushing syndrome from cynicism would be mediated by these corticosteroid pathways acting on modified calcium (hsa04020) and cAMP (hsa04024) signaling. Our results here suggest a testable hypothesis to relate cynicism to Cushing syndrome and creating a potential intervention by where reducing occupational burnout could lead to fewer adrenal disorders.

The presence of genetic markers associated with increased cynicism and CD may be useful in identifying individuals who may be at a higher risk for negative health outcomes. This could allow for early intervention and prevention strategies to be put in place to reduce the risk of these negative outcomes. For example, practicing mindfulness and gratitude are recognized methods for combating cynicism [28, 29] and may help improve heart health [30]. Research suggests that a regular meditation practice could protect against heart disease by improving heart rate, reducing stress and lowering blood pressure. Mindfulness meditation may be useful in reducing cynicism [23], resulting in handling stress in a healthy manner and improving overall well-being and quality of life.

## CONCLUSION

Here we reported our results from a GWAS for cynicism and CD. Using different inheritance models and accounting for population stratification, we identified multiple significant associations with each trait and five associations shared between the traits. We also performed an unadjusted GWAS in an attempt to reproduce previously published results using a two-stage discovery and validation approach. Using the same underlying dataset, we were able to reproduce 17% of the significant variants. Taken together, the results of the present study reinforce the implications of the first study that there appears to be a significant genetic component to the development of cynicism and CD throughout a lifetime.

## Supporting information

Supplemental Table 1 - Significant Variants

Supplementary Figure S1 - PC Variance Explained

Supplementary Figure S2 - PCA Analysis

## Data Availability

Genotype and phenotype data that support this study can be obtained from dbGAP using accession numbers phs000285.v3.p2, pht001579.v2.p2, pht001634v2.p2, pht001786.v2.p2, phg000092.v2, and phg000098.v2. GWAS summary statistics are unavailable for public posting, per the CARDIA data use agreement. The lists of significantly associated variants along with P-value and effect sizes are available as supplementary material. Full summary statistics are available from the author upon reasonable request.

## ACKNOWLEDGMENTS

The authors wish to thank Mr. Billy Thompson, Ms. CharLee Martin, Mr. Eric Rigby, and Mr. Warren Fridy for assistance in data access and security. The authors would also like to thank Dr. Tyler Mulhearn and Ms. Tanya Goodman for their critical comments and review. The authors also wish to thank the participants and the original depositors of the CARDIA study.

## Footnotes

### Institutional review board statement

This study was reviewed and approved by WCG Institutional Review Board (Study number 1332892).

### Data sharing statement

The authors have read the STREGA guidelines for reporting genetic association studies and prepared the manuscript accordingly.

