## Supplementary figures and images for "Toward Understanding Genetic Risk of Burnout through Genome-wide Associations of Cynicism and Cynical Distrust"

### Supplementary Figure S1 - PC Variance Explained

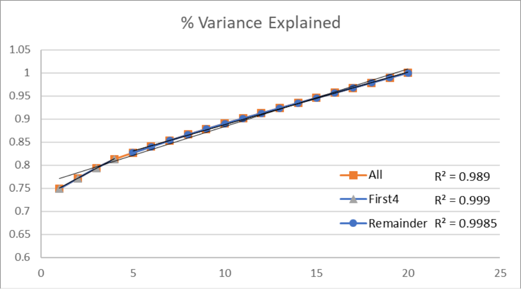

### Supplementary Figure S2 - PCA Analysis

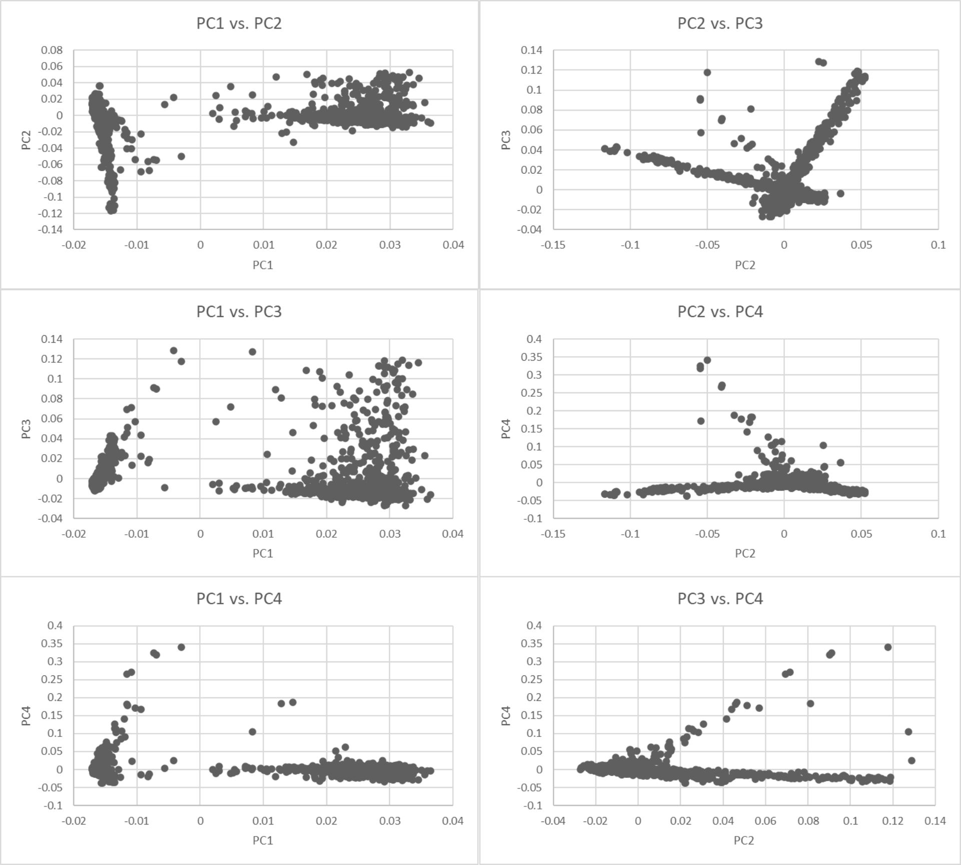
